# Influence of a Large Language Model on Diagnostic Reasoning: A Randomized Clinical Vignette Study

**DOI:** 10.1101/2024.03.12.24303785

**Authors:** Ethan Goh, Robert Gallo, Jason Hom, Eric Strong, Yingjie Weng, Hannah Kerman, Josephine Cool, Zahir Kanjee, Andrew S. Parsons, Neera Ahuja, Eric Horvitz, Daniel Yang, Arnold Milstein, Andrew P.J Olson, Adam Rodman, Jonathan H Chen

## Abstract

**Importance:** Diagnostic errors are common and cause significant morbidity. Large language models (LLMs) have shown promise in their performance on both multiple-choice and open-ended medical reasoning examinations, but it remains unknown whether the use of such tools improves diagnostic reasoning.

**Objective:** To assess the impact of the GPT-4 LLM on physicians’ diagnostic reasoning compared to conventional resources.

**Design:** Multi-center, randomized clinical vignette study.

**Setting:** The study was conducted using remote video conferencing with physicians across the country and in-person participation across multiple academic medical institutions.

**Participants:** Resident and attending physicians with training in family medicine, internal medicine, or emergency medicine.

**Intervention(s):** Participants were randomized to access GPT-4 in addition to conventional diagnostic resources or to just conventional resources. They were allocated 60 minutes to review up to six clinical vignettes adapted from established diagnostic reasoning exams.

**Main Outcome(s) and Measure(s):** The primary outcome was diagnostic performance based on differential diagnosis accuracy, appropriateness of supporting and opposing factors, and next diagnostic evaluation steps. Secondary outcomes included time spent per case and final diagnosis.

**Results:** 50 physicians (26 attendings, 24 residents) participated, with an average of 5.2 cases completed per participant. The median diagnostic reasoning score per case was 76.3 percent (IQR 65.8 to 86.8) for the GPT-4 group and 73.7 percent (IQR 63.2 to 84.2) for the conventional resources group, with an adjusted difference of 1.6 percentage points (95% CI -4.4 to 7.6; p=0.60). The median time spent on cases for the GPT-4 group was 519 seconds (IQR 371 to 668 seconds), compared to 565 seconds (IQR 456 to 788 seconds) for the conventional resources group, with a time difference of -82 seconds (95% CI -195 to 31; p=0.20). GPT-4 alone scored 15.5 percentage points (95% CI 1.5 to 29, p=0.03) higher than the conventional resources group.

**Conclusions and Relevance:** In a clinical vignette-based study, the availability of GPT-4 to physicians as a diagnostic aid did not significantly improve clinical reasoning compared to conventional resources, although it may improve components of clinical reasoning such as efficiency. GPT-4 alone demonstrated higher performance than both physician groups, suggesting opportunities for further improvement in physician-AI collaboration in clinical practice.

## INTRODUCTION

Medical diagnosis is a high-stakes cognitive process that takes place in time-constrained and stressful clinical environments. Diagnostic errors are common and contribute to significant patient harm^1,2,3,4,5,6^. Strategies to reduce diagnostic errors include a variety of educational, reflective, and team-based practices. The impacts of these interventions have been limited, and even the most effective methods are difficult to integrate into clinical practice at scale^7,8^. Artificial intelligence (AI) technologies have long been pursued as promising tools for assisting physicians with diagnostic reasoning. To date, research on AI in medicine has largely focused on diagnosis and prediction of outcomes in specific domains.

New technological improvements in large language models (LLMs) – machine learning systems that produce human-like responses from free text prompts – have shown the ability to solve complex cases, display human-like clinical reasoning, take patient histories, and communicate empathetically^9,10,11,12,13,14^. LLMs can be scaled into a variety of clinical workflows given their generalizable nature, and are already being integrated into healthcare^15,16^. Early integrations of LLMs will almost certainly require a “human in the loop” – augmenting, rather than replacing, human expertise and oversight^17^. Despite the impressive performance of these emerging technologies in experimental settings and rapid moves toward integration into clinical practice, considerable gaps remain in our understanding of how these systems affect human performance. Meaningful measures of the quality of diagnostic reasoning may help close this gap.

We performed a randomized clinical vignette study using complex diagnostic cases to compare the diagnostic reasoning performance of physicians using a commercial AI chatbot (ChatGPT Plus, GPT-4) with the performance of physicians using conventional diagnostic reference resources. To move beyond simplistic evaluations of diagnostic accuracy, we further developed and validated a novel assessment tool adapted from the literature on human diagnostic reasoning, structured reflection^18^.

## METHODS

**Figure 1:**
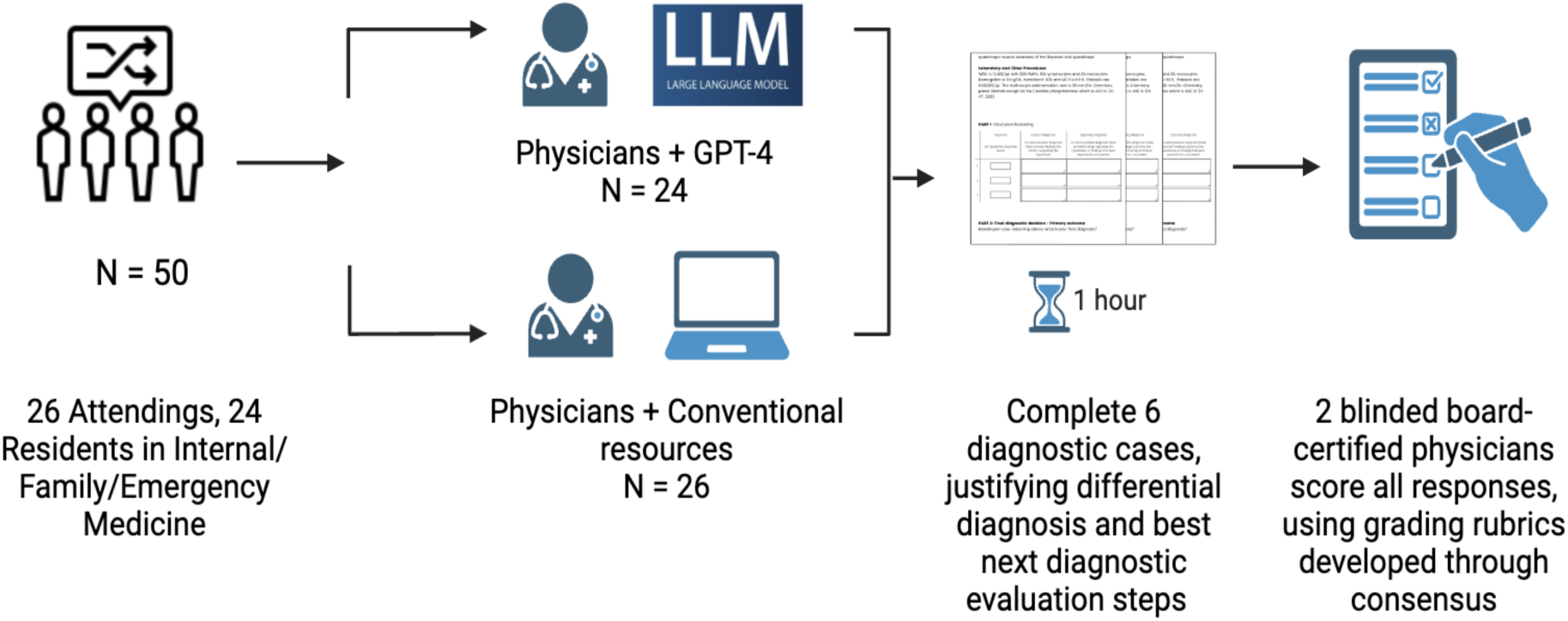
50 physicians randomized to complete diagnosis quiz with GPT-4 vs. conventional resources. Participants were asked to offer differential diagnosis with supporting statements of findings in favour or against each differential, and to propose best next diagnostic evaluation steps.

We recruited practicing attendings and residents with training in a general medical specialty (internal medicine, family medicine, or emergency medicine) through email lists used for community messaging at Stanford University, Beth Israel Deaconess Medical Center, and the University of Virginia. Informed consent was obtained prior to enrollment and randomization. Small groups of participants were proctored by study coordinators either remotely or at an in-person computer laboratory. Sessions lasted for one hour. Resident participants were offered $100 and attending participants were offered up to $200 for completing the study.

### Clinical Vignettes

Clinical vignettes were adapted from a landmark study that set the standards for the evaluation of computer-based diagnostic systems, including developing measures of diagnostic accuracy and relevance^19^. All cases were based on actual patients and included data available on initial diagnostic evaluation, including history, physical exam, and results of laboratory tests. The cases have never been publicly released to protect the validity of the test materials for future use; therefore, it is unlikely that the materials are included in GPT-4’s training data. Figure 2 includes a representative example of one of the cases. After iterative discussion among the investigators of all 110 cases, 6 were chosen to reflect diagnostic challenges across different adult medicine specialties. Cases were edited to reflect modern laboratory evaluation (e.g., referring to AST rather than SGOT) as necessary and pilot-tested with two groups of participants not in the study.

A common gold standard in clinical decision support diagnostic studies has been the accuracy of differential diagnosis. Methods for the assessment of clinical reasoning by humans are far richer and include a variety of strategies including objective structured clinical exams (OSCEs), script concordance testing, evaluation of documentation, and global assessments adapted from the psychological literature^20^. To better capture the richness and nuance of diagnostic reasoning, we treated diagnostic accuracy as a secondary outcome, and instead developed and validated as a primary measure of performance a more holistic assessment of reasoning, which we refer to as structured reflection.

Structured reflection is aimed at capturing and improving the process by which physicians consider reasonable diagnoses and case features that support or oppose their diagnoses, similar to how physicians may explain their reasoning in the “Assessment and Plan” component of clinical notes^21,22^. Adapting previous methodologies demonstrated to improve diagnostic performance, participants completed a structured reflection grid with free text responses. After user testing, we simplified the grid by collapsing two categories – evidential features that were missing and features that would have been expected but were not present – into a single category of features opposing the diagnosis. Additionally, participants were asked to provide their most likely diagnosis and up to three next steps to further evaluate the patient.

### Grading of Performance

We built upon previous studies of structured reflection by scoring the rubric itself, not just final diagnosis accuracy. For each case, we assigned up to 1 point for each plausible diagnosis. Findings supporting each diagnosis and findings opposing the diagnosis were also graded based on correctness, with 0 points for incorrect or absent answers, 1 point for partially correct, and 2 points for completely correct responses. The final diagnosis was graded as 2 points for the most correct diagnosis, while 1 point was awarded for a plausible diagnosis or a correct diagnosis that was not specific enough compared to the most correct diagnosis. Finally, participants were instructed to describe up to 3 next steps to further evaluate the patient with 0 points awarded for an incorrect response, 1 point awarded for a partially correct response and 2 points for a completely correct response (see Supplementary 2, eTable 2). Participants who had incorrect differential diagnosis items but reasonable reasoning based on those items were not penalized.

### Study Design

We employed a randomized, single-blinded study design. Participants were randomized to access GPT-4 via the ChatGPT Plus interface (intervention group) or to conventional resources only (control group). Both groups were permitted to access any resources they normally use for clinical care (with examples given of UpToDate [Wolters Kluwer, Philadelphia, PA], Epocrates [Athenahealth, Watertown, MA], and Google [Google, Mountain View, CA] search); the control group was explicitly instructed not to use large language models (e.g., ChatGPT, Bard, Claude, MedPaLM, LLAMA2, etc.). Participants had one hour to complete as many of the six diagnostic cases as they could. Participants were instructed to prioritize quality of responses over completing all cases.

The study was conducted using a Qualtrics survey tool. Each case presented a clinical vignette for which participants were asked to complete the structured reflection process described above. Cases were presented in random order for each participant. In a secondary analysis, we included a comparison arm using GPT-4 alone to answer the cases. Using established principles of prompt design, we iteratively developed a few-shot prompt – a type of input where the language model is given examples to follow – by copy-pasting the clinical vignette questions^23^. For the prompt, we used the same example provided to the human participants (Supplementary 5, eTable 5). These were run three times, and the results from the three runs were included for blinded grading alongside the human outputs before any unblinding or data analysis.

### Assessment Tool Validation

In order to establish validity in our population, we collected two sets of data which were not included in the final study, with 13 participants in total. The three primary scorers (J.H, A.R, and A.O.), all board-certified physicians with experience in the evaluation of clinical reasoning at the post-graduate medical level, graded each of these sets together, to ensure consistency. After data collection, each case was graded independently by 2 scorers who were blinded to the assigned treatment group. When scorers disagreed, they met to engage on the differences in their assessments and to seek consensus. Disagreement was predefined by a difference of > 10% of the final score, based on experience that this represented a clinically significant disagreement. In addition, the final diagnosis scoring was adjudicated by two reviewers to obtain agreement for the secondary outcome of diagnostic accuracy. We calculated a weighted Cohen’s kappa to show concordance in grading. We calculated Cronbach’s alpha to determine the internal reliability of this measure.

### Study outcome

Our primary outcome was the final score as a percentage across all components of the structured reflection tool. A key secondary outcome was time spent per case in seconds. Final diagnosis accuracy, a common primary outcome in diagnosis studies, was evaluated as a secondary outcome. Final diagnosis was treated as an ordinal outcome with three groups (incorrect, partially correct, and most correct). Since the difference between the most correct response and partially correct responses may not be clinically meaningful, we additionally analyzed the outcomes as binary (incorrect compared to at least partially correct). This represents the avoidance of a diagnostic error.

### Statistical Analysis

The target sample size of 50 participants was pre-specified based on a power analysis using our two validations sets of data, scored prior to study enrollment, corresponding to an expected 200 to 250 cases completed (4-5 cases per participant). All analyses were at the case level, clustered by the participant. In the primary analysis, we only included cases with completed responses. Generalized mixed-effect models were applied to assess the difference in the primary and secondary outcomes of the GPT-4 group compared to the conventional resources only group. A random-effect for the participant was included in the model to account for the potential correlation between cases for a participant. Additionally, a random effect for cases was included to account for any potential variability in difficulty across cases. A pre-planned sensitivity analysis evaluated the effect of including incomplete cases on the primary outcome. Subgroup analyses were conducted based on training status and experience with ChatGPT. In a secondary analysis, cases completed by GPT-4 alone were treated as a third group with cases clustered in a nested structure of 3 attempts under a single participant. These were compared to cases from real participants with each case considered as a single attempt under a single participant using a similar nested structure. All statistical analysis was performed using R v4.3.2 (R Foundation for Statistical Computing, Vienna, Austria). Statistical significance was based on a p value <0.05. This study was reviewed and determined to be exempt by institutional review boards at Stanford University, Beth Israel Deaconess Medical Center, and University of Virginia.

## RESULTS

50 US-licensed physicians were enlisted (26 attendings, 24 residents). Median years in practice was 3 (IQR 2-9). Further information on participants is included in Table 1 below.

**Table 1:**
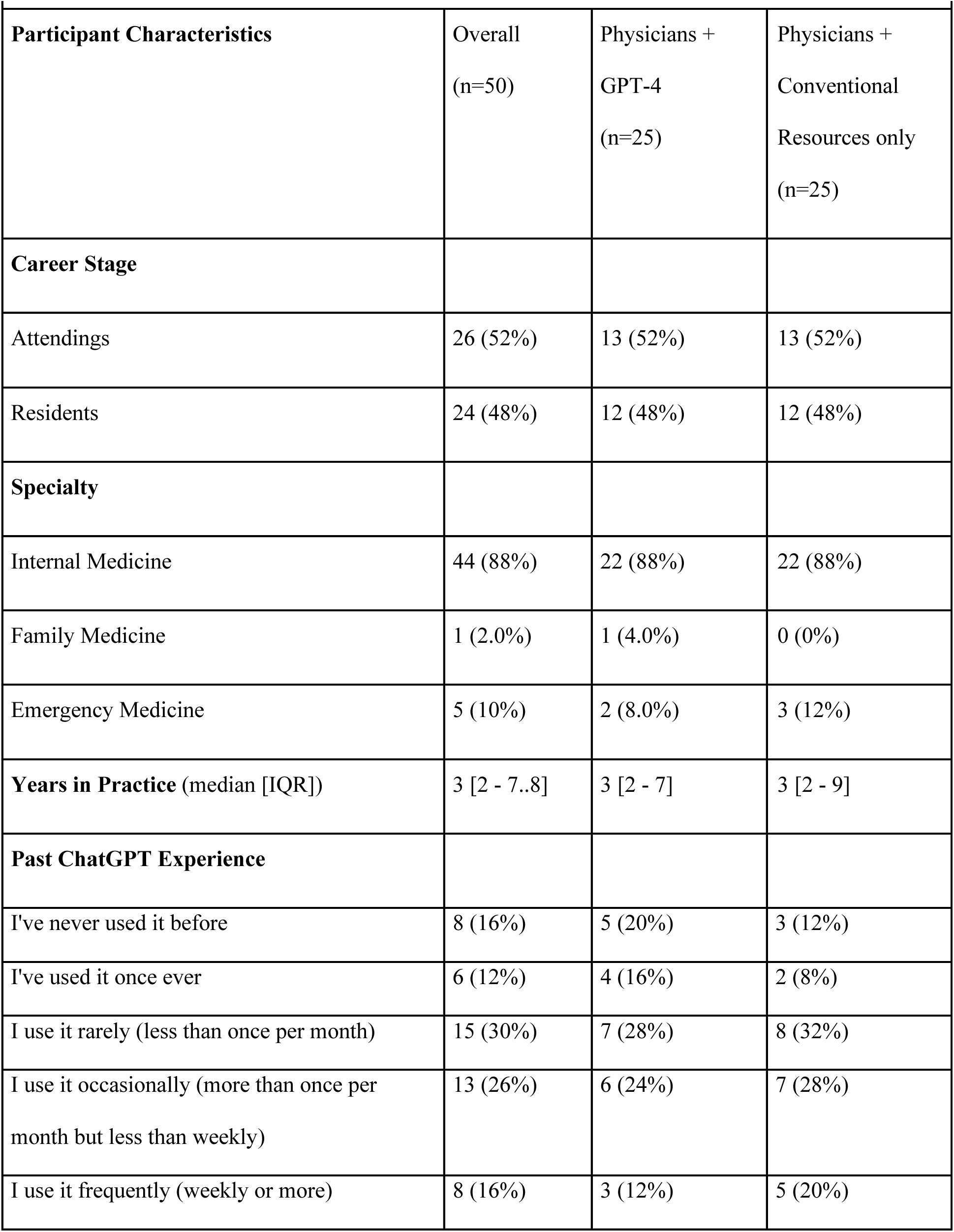
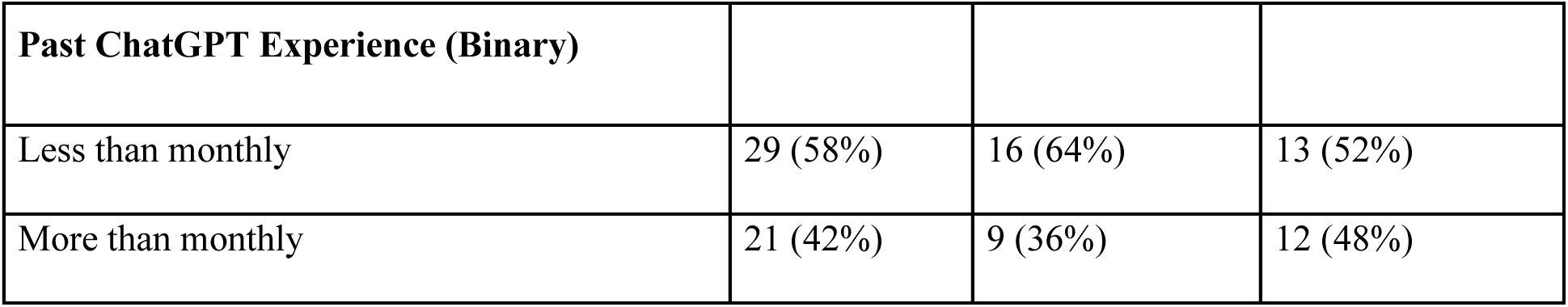
Baseline Participant Characteristics.

| **Table 1: Baseline Participant Characteristics** |  |  |  |
| --- | --- | --- | --- |
| **Participant Characteristics** | Overall<br>(n=50) | Physicians +<br>GPT-4<br>(n=25) | Physicians +<br>Conventional<br>Resources only<br>(n=25) |
| **Career Stage** |  |  |  |
| Attendings | 26 (52%) | 13 (52%) | 13 (52%) |
| Residents | 24 (48%) | 12 (48%) | 12 (48%) |
| **Specialty** |  |  |  |
| Internal Medicine | 44 (88%) | 22 (88%) | 22 (88%) |
| Family Medicine | 1 (2.0%) | 1 (4.0%) | 0 (0%) |
| Emergency Medicine | 5 (10%) | 2 (8.0%) | 3 (12%) |
| **Years in Practice** (median [IQR]) | 3 [2 - 7..8] | 3 [2 - 7] | 3 [2 - 9] |
| **Past ChatGPT Experience** |  |  |  |
| I've never used it before | 8 (16%) | 5 (20%) | 3 (12%) |
| I've used it once ever | 6 (12%) | 4 (16%) | 2 (8%) |
| I use it rarely (less than once per month) | 15 (30%) | 7 (28%) | 8 (32%) |
| I use it occasionally (more than once per month but less than weekly) | 13 (26%) | 6 (24%) | 7 (28%) |
| I use it frequently (weekly or more) | 8 (16%) | 3 (12%) | 5 (20%) |

| Past ChatGPT Experience (Binary) |  |  |  |
| --- | --- | --- | --- |
| Less than monthly | 29 (58%) | 16 (64%) | 13 (52%) |
| More than monthly | 21 (42%) | 9 (36%) | 12 (48%) |

### Primary Outcome: Diagnostic performance

Median number of completed cases was 5.2. The median score per case was 76.3 (IQR 65.8 to 86.8) for the GPT-4 group and 73.7 (IQR 63.2 to 84.2) for the conventional resources group. The generalized mixed effects model resulted in a difference of 1.6 percentage points (95% CI -4.4, 7.6; p=0.6) between the GPT-4 and conventional resources groups as shown in Table 2. A sensitivity analysis including all cases, complete and incomplete, showed a similar result with a difference of 2.0 percentage points (95% CI -4.1 to 8.2; p=0.5) between the GPT-4 and conventional resources group.

**Table 2:** Performance Outcomes.

| **Table 2: Performance Outcomes** |  |  |  |  |
| --- | --- | --- | --- | --- |
| Group | Diagnostic Performance (percentage points) |  |  |  |
|  | Physicians + GPT-4 | Physicians +<br>Conventional Resources | Difference<br>(95%CI) | P-value |
| All<br>Participants | 76.3 (65.8, 86.8) | 73.7 (63.2, 84.2) | 1.6 (-4.4, 7.6) | 0.60 |
| **Level of Training** |  |  |  |  |
| Attending | 78.9 (63.2, 86.8) | 75.0 (60.5, 86.8) | 0.5 (-8.9, 9.9) | 0.92 |
| Resident | 76.3 (68.4, 84.2) | 73.7 (63.2, 84.2) | 2.8 (-5.5, 11.1) | 0.50 |
| **ChatGPT Experience** |  |  |  |  |
| Less than<br>monthly | 76.3 (63.2, 84.2) | 76.3 (63.2, 86.8) | -0.5 (-7.7, 6.7) | 0.90 |
| More than<br>monthly | 78.9 (67.8, 89.5) | 73.7 (62.5, 84.2) | 4.5 (-6.7, 15.7) | 0.40 |

*Caption: Continuous variables are expressed as median (interquartile range). Differences between groups are reported from the multilevel analysis accounting for clustering of cases by participant.*

### Secondary Outcomes

The median time spent per case was 519 seconds (IQR 371 to 668 seconds) for the GPT-4 group and 565 seconds (IQR 456 to 788 seconds) for the conventional resources group (Table 3). The linear mixed effects model resulted in an adjusted difference of -82 seconds (95% CI -195 seconds to 31 seconds; p=0.20).

**Table 3:**
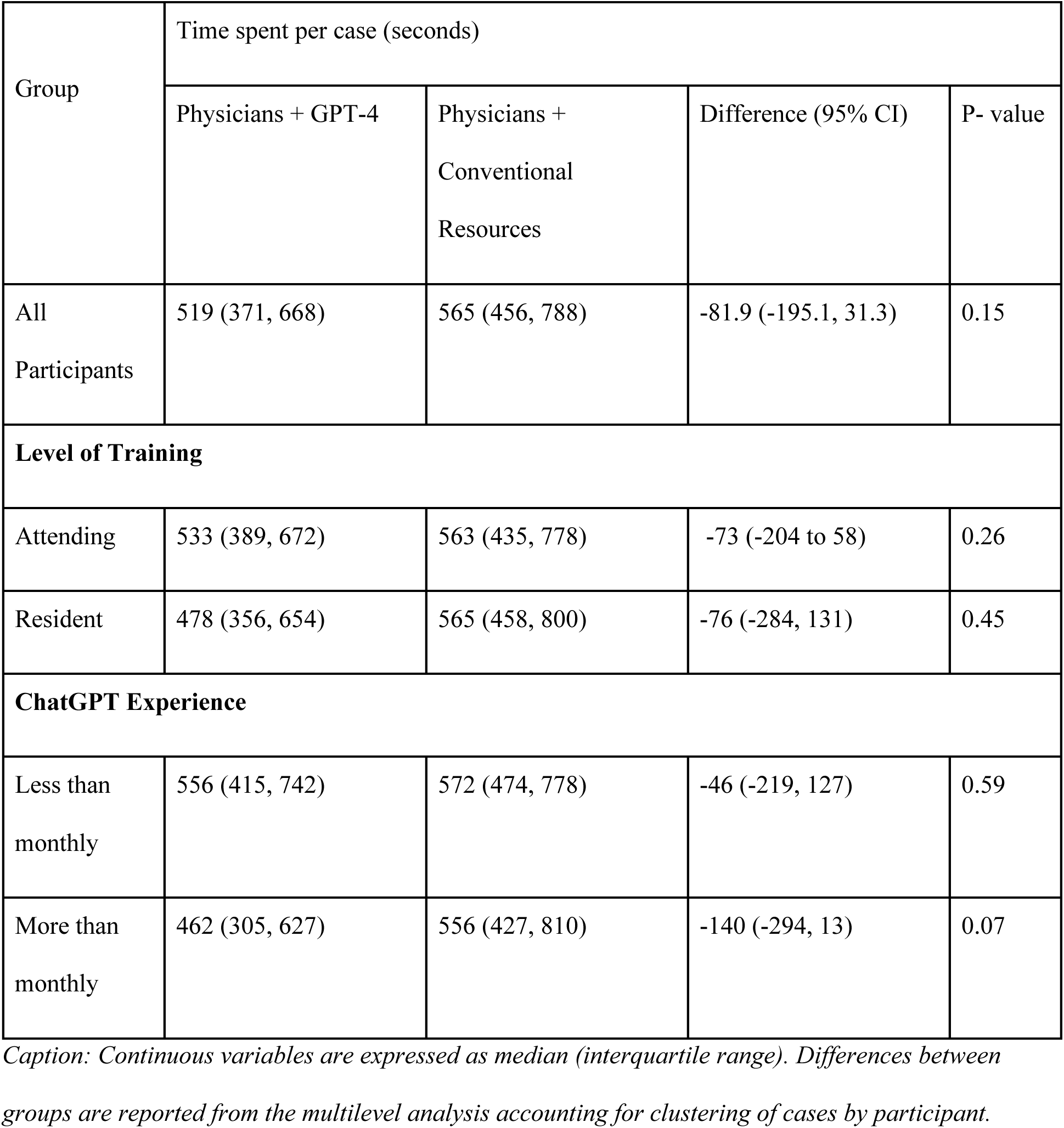
Time Spent Per Case.

Accuracy of final diagnosis was evaluated as well, as shown in Supplementary 3, eTable 3. Using the ordinal scale, the GPT-4 group had a 1.4 higher odds (95% CI 0.67 to 2.8; p=0.39) of a more correct diagnosis. Treating final diagnosis as binary correct compared to incorrect did not qualitatively change the results (OR 1.9, 95% CI 0.9 to 4.0; p=0.10).

### Subgroup Analyses

Tables 2 and 3 include the analyses by subgroups, including level of training and level of prior experience with chatGPT. Subgroup analyses were directionally similar to the analyses for the whole cohort.

### GPT-4 alone

In the three runs of GPT-4 alone, the median score per case was 92.1 percentage points (IQR 82.2 to 97.4). Comparing GPT-4 *alone* to the human with conventional resources group found a score difference of 15.5 percentage points (95% CI 1.5 to 29.5 percentage points; p=0.03) favoring GPT-4 alone (Supplementary 4, eTable 4).

### Assessment Tool Validation

The weighted Cohen’s kappa between all three graders was 0.66, indicating substantial agreement within the expected range for diagnostic performance studies^24^. The overall Cronbach’s alpha was 0.64. The variances of individual sections of the structured reflection rubric are shown in Supplementary 6, eTable 6. After removing Final Diagnosis, which had the highest variance, the Cronbach’s alpha was 0.67.

## DISCUSSION

This randomized clinical vignette study found that physician use of a commercially available LLM chatbot did not improve diagnostic reasoning on challenging clinical cases, despite the LLM alone outperforming human participants. The results were similar across the important subgroups of different training levels and experience with the chatbot. Since the task in this study is similar to how physicians often structure their clinical assessments and plans, these results suggest that providing access to GPT-4 alone may not improve overall diagnostic reasoning in clinical practice. These findings are particularly relevant now that many health systems offer HIPAA-compliant chatbots that physicians can use for clinical care^25^.

Even though we did not find a meaningful difference in diagnostic reasoning overall with access to GPT-4, the LLM may improve physician performance in certain areas of clinical reasoning. The average time spent on cases for those randomized to the GPT-4 arm was almost a minute less per case and over two minutes less per case for the subgroup who reported occasional or frequent use of the chatbot. Given the wide variability in time to complete cases, the results for time spent per case did not reach statistical significance despite suggesting a potentially relevant difference. Final diagnosis accuracy also potentially included a meaningful benefit, but this outcome did not reach statistical significance either.

If confirmed with additional studies, improvement in diagnostic efficiency and final diagnosis accuracy may be enough to justify the use of LLM chatbots in clinical practice given the time-constrained nature of clinical medicine and the need to address the long-term challenge of diagnostic error^26^. An important barrier to the use of clinical decision support systems in medicine is the integration into clinical workflows without increasing physician workload and time spent in the electronic health record; if LLMs are able to increase efficiency without sacrificing performance, then they may prove well worth the cost to securely house the models and train physicians in their clinical application.

A surprising result of a secondary analysis was that the LLM alone performed significantly better on diagnostic challenges than both groups of humans, which is consistent with a prior study^27^. These results should not be interpreted to mean that LLMs should be used for diagnosis without physician oversight. Our study and others were performed using clinical case vignettes that were curated and summarized by human clinicians with specific and answerable diagnostic questions in mind and do not reflect the full ambiguity in patient care settings. These vignette-style cases address an important, but specific component of diagnostic reasoning – the ability to extract both relevant and exculpatory information from case vignettes with relatively little “noise”. While early studies show that LLMs might effectively collect and summarize patient information, these capabilities will need to be explored more thoroughly^27,28^.

The difference between the performance of the LLM alone and that of the clinicians provided with access to the LLM highlights important opportunities for research on enhanced human-clinician collaboration. For one, studies have demonstrated that the accuracy of LLM output is sensitive to the formulation of prompts, and therefore prompt engineering by the study team could explain the superior performance of GPT-4 alone compared to the study participants^29^. Training clinicians in best prompting practices may improve physician performance with LLMs. Alternatively, predefined “prompting for diagnostic decision support” might be optimized as a system service for physicians. Second, we note the rich design space for exploring and enhancing clinician-AI interaction, including gaining better understandings of how and when to display AI inferences to physicians^30^. Our results highlight the potential for improving the diagnostic performance of physicians through innovation with integrating AI capabilities into clinical workflows. More generally, we see opportunity with deliberate consideration and redesign of medical education and practice frameworks that enable the best use of computer and human resources to deliver optimal medical care.

Our study also developed and validated a measure, structured reflection, inspired by studies of physician cognition. This assessment tool demonstrated substantial agreement between graders and internal reliability similar or superior to other measures used in the assessment of reasoning^31,32,33,34^. Early research focused on benchmarks with limited clinical utility, such as multiple-choice question banks used for medical licensing; or curated case vignettes of diseases rarely seen in clinical practice such as *New England Journal of Medicine* clinicopathological case conferences^11,35^. While having obvious advantages in ease of measurement, these tasks are not consistent with clinical reasoning in practice. We must understand how AI affects reasoning for implementation purposes, rather than merely demonstrating an improvement in multiple choice answering or diagnoses rarely encountered in clinical practice. As AI systems become more advanced and autonomous, we must urgently ensure their alignment with human needs and thought processes.

### Limitations

We focused our investigation around a single LLM, GPT-4, given its commercial availability and integration into clinical practice^25^. Multiple alternative LLM systems are rapidly emerging, though GPT-4 currently remains amongst the most performant for the applications studied^36,37^. Participants were given access to the GPT-4 chatbot without explicit training in prompt engineering techniques that could have improved the quality of their interactions with the system, however this is consistent with many current integrations^25^. Our cohort was a convenience sample from multiple major academic centers, so our results may not be representative of the broader population of practicing physicians. Our study included six cases that were deliberately selected to ensure a broad and relevant selection of medicine cases, but any sample could never cover the full variety of cases to represent the field of medicine. Our approach and total number of cases is nonetheless consistent with established human criterion-based assessments, including national licensing assessments in which students completed 12 cases over 8 hours^38,39,40,41,42^. Given the internal reliability of our assessment, there is no evidence to suggest that additional case sampling would meaningfully alter the overall results of this study.

## CONCLUSION

Despite GPT-4 alone significantly outscoring human physicians on a complex diagnostic reasoning clinical vignette study, the availability of GPT-4 as a diagnostic aid did not improve physician performance compared to conventional resources. While the use of a large language model may improve the correctness of final diagnosis and efficiency of diagnostic reasoning, further development is needed to effectively integrate AI into emerging clinical decision support systems to exploit their potential for improving medical diagnosis in practice.

### Data Availability

Example case vignettes, questions, and grading rubrics are included in the supplement. GPT-4 transcript chat logs, raw score table, and individual survey responses are available upon request.

## CONTRIBUTIONS

Ethan Goh (co-first author) - Study design, data acquisition, data interpretation, manuscript preparation

Robert Gallo (co-first author) - Study design, data acquisition, data interpretation, manuscript preparation

Jason Hom - Study design, data acquisition, data interpretation

Eric Strong Study design, data acquisition, data interpretation

Yingjie Weng - Study design, data interpretation

Hannah Kerman - Study design, data interpretation

Josephine Cool - Study design, data interpretation

Zahir Kanjee - Study design, data interpretation

Andrew Parsons - Study design, data interpretation

Daniel Yang - Study design, data interpretation

Arnold Milstein - Funding and administrative support

Neera Ahuja - Funding and administrative support

Eric Horvitz - Study design, data interpretation

Andrew Olson (co-last author) - Study design, data analysis, data interpretation, critical revision, supervision

Adam Rodman (co-last author) - Study design, data analysis, data interpretation, critical revision, supervision

Jonathan Chen (co-last author) - Study design, data analysis, data interpretation, critical revision, supervision, funding and administrative support

## AFFILIATIONS, DISCLOSURES AND FUNDING

### Ethan Goh, MD, MS

#### Affiliations

- Stanford Center for Biomedical Informatics Research, Stanford University, Stanford, California, USA
- Stanford Clinical Excellence Research Center, Stanford University, Stanford, California, USA

#### Disclosures

- None

#### Funding

- Gordon and Betty Moore Foundation

### Robert Gallo, MD

#### Affiliations

- Center for Innovation to Implementation, VA Palo Alto Health Care System

#### Disclosures

- None

#### Funding

- Dr. Gallo is supported by a VA Advanced Fellowship in Medical Informatics. The views expressed are those of the authors and not necessarily those of the Department of Veterans Affairs or those of the United States government.

### Jason Hom, MD

#### Affiliations

- Stanford University School of Medicine

#### Disclosures

- None

#### Funding

- Gordon and Betty Moore Foundation (Grant #12409)

### Eric Strong, MD

#### Affiliations

- Stanford University School of Medicine

#### Disclosures

- None

#### Funding

- Gordon and Betty Moore Foundation (Grant #12409)

### Yingjie Weng, MHS

#### Affiliations

- Quantitative Sciences Unit, Stanford University School of Medicine, Palo Alto, CA

#### Disclosures

- None

#### Funding

- None

### Hannah Kerman, MD

#### Affiliations

- Beth Israel Deaconess Medical Center, Boston, MA
- Harvard Medical School, Boston, MA

#### Disclosures

- None

#### Funding

- None

### Josephine Cool, MD

#### Affiliations

- Beth Israel Deaconess Medical Center, Boston, MA
- Harvard Medical School, Boston, MA

#### Disclosures

- None

#### Funding

- Gordon and Betty Moore Foundation

### Zahir Kanjee, MD, MPH

#### Affiliations

- Beth Israel Deaconess Medical Center, Boston, MA
- Harvard Medical School, Boston, MA

#### Disclosures

- Royalties from Wolters Kluwer for books edited (unrelated to this study), former paid advisory member for Wolters Kluwer on medical education products (unrelated to this study), honoraria from Oakstone Publishing for CME delivered (unrelated to this study)

#### Funding

- Gordon and Betty Moore Foundation

### Andrew S. Parsons, MD, MPH

#### Affiliations

- University of Virginia School of Medicine

#### Disclosures

- Paid advisory role for New England Journal of Medicine (NEJM) Group and National Board of Medical Examiners (NBME) for medical education products (unrelated to this study)

#### Funding

- None

### Neera Ahuja, MD

#### Affiliations

- Stanford University School of Medicine

#### Disclosures

- None

#### Funding

- None

### Eric Horvitz, PhD, MD

#### Affiliations

- Microsoft
- Stanford HAI

#### Disclosures

- None

#### Funding

- None

### Andrew P.J. Olson, MD

#### Affiliations

- University of Minnesota Medical School, Minneapolis, Minnesota
- Division of Hospital Medicine, Department of Medicine
- Division of Pediatric Hospital Medicine, Department of Pediatrics

#### Disclosures

- Dr. Olson receives funding from 3M for research related to rural health workforce shortages. Dr. Olson receives consulting fees for work related to a clinical reasoning application from the New England Journal of Medicine.

#### Funding

- Gordon and Betty Moore Foundation

### Adam Rodman, MD, MPH

#### Affiliations

- Beth Israel Deaconess Medical Center, Boston, MA.
- Harvard Medical School, Boston, MA

#### Disclosures

- None

#### Funding

- Gordon and Betty Moore Foundation

### Daniel Yang, MD

#### Affiliations

- Kaiser Permanente, Oakland, CA

#### Disclosures

- None Funding
- None

### Arnold Milstein, MD

Affiliations

- Stanford Clinical Excellence Research Center, Stanford University, Stanford, California, USA

Disclosures

- Dr Milstein reported uncompensated and compensated relationships with care.coach, Emsana Health, Embold Health, EZPT, FN Advisors, Intermountain Healthcare, JRSL, The Leapfrog Group, Peterson Center on Healthcare, Prealize Health, PBGH

## Funding

- Pooled philanthropic gifts to Stanford University
- Research funding from Stanford Healthcare and Stanford Children’s Health

### Jonathan H. Chen

#### Affiliations

- Stanford Center for Biomedical Informatics Research, Stanford University, Stanford, California, USA
- Division of Hospital Medicine, Stanford University, Stanford, California, USA
- Stanford Clinical Excellence Research Center, Stanford University, Stanford, California, USA

#### Disclosures

- Co-founder of Reaction Explorer LLC that develops and licenses organic chemistry education software.
- Paid consulting fees from Sutton Pierce, Younker Hyde MacFarlane, and Sykes McAllister as a medical expert witness.

#### Funding

- NIH/National Institute of Allergy and Infectious Diseases (1R01AI17812101)
- NIH/National Institute on Drug Abuse Clinical Trials Network (UG1DA015815 - CTN-0136)
- Gordon and Betty Moore Foundation (Grant #12409)
- Stanford Artificial Intelligence in Medicine and Imaging - Human-Centered Artificial Intelligence (AIMI-HAI) Partnership Grant
- Doris Duke Charitable Foundation - Covid-19 Fund to Retain Clinical Scientists (20211260)
- Google, Inc. Research collaboration Co-I to leverage EHR data to predict a range of clinical outcomes.
- American Heart Association - Strategically Focused Research Network - Diversity in Clinical Trials

## Data Availability

All data produced in the present study are available upon reasonable request to the authors

