## Supplementary Materials for "Influence of a Large Language Model on Diagnostic Reasoning: A Randomized Clinical Vignette Study"

- Supplementary 1: Clinical Vignette
- Supplementary 2: Grading Rubric
- Supplementary 3: Distribution of Diagnostic Performance Scores
- Supplementary 4: GPT-4 Performance
- Supplementary 5: GPT Prompt and responses
- Supplementary 6: Assessment Tool Validation

**SUPPLEMENTARY 1: CLINICAL VIGNETTE**

**eTable 1: Diagnostic Case #1 Vignette and Questions**

| **Diagnostic Vignette** | |
| --- | --- |
| **History Of Present Illness** A 76M comes to his PCP complaining of pain in his back and thighs for 2 weeks. He has no pain sitting or lying, but walking causes severe pain in his low back, buttocks and calves. He feels febrile and tired. He was told by the referring cardiologist that his recent tests results since the pain started showed a new anemia and azotemia. A few days before the onset of the pain he had undergone coronary angioplasty. Heparin was administered for 48 hours.  **Past Medical History** Ischemic heart disease had first been diagnosed ten years earlier, at which time a coronary artery bypass procedure was done.  **Physical Examination** VITALS: 99.6° F.; pulse was 94/min and regular; BP was 110/88 mmHg. GEN: Well appearing CARDS: There is a grade III/VI apical systolic murmur. PULM: Lungs are clear to auscultation bilaterally, no wheezing, or consolidations noted ABD: Soft, non-tender to palpation MSK: He does not have tenderness of his spine or pelvis. Spinal mobility is normal, as is the mobility of his hips. Standing is painless; however, pain is experienced in his low back, buttocks and calves within a minute of feeble running in place. The pain disappears shortly after exercise is discontinued. EXT: Peripheral pulses were symmetrically reduced, but palpable. The neurological examination was normal. SKIN: Patient has a purple, red, lacy rash over his low back and buttocks.  **Laboratory** WBC of 11.5 x 103 cells /μL; differential of 64% segs, 20% lymphocytes, 3% monocytes, 12% eosinophils and 1% basophil. The hematocrit was 28% and the platelet count was 315 x 103 /μL. The erythrocyte sedimentation rate was 99 mm/h. Urinalysis was normal except for 2+ proteinuria. Serum creatinine was 4.0 mg/dL; sodium was 145 mEq/L; potassium 4.0 mEq/L; chloride 105 mEq/L. SGOT was 27 U/L; GGT was 90 U/L; alkaline phosphatase was 153 U/L. | |
| **Part 1 – Structured Reasoning Grid** | |
| 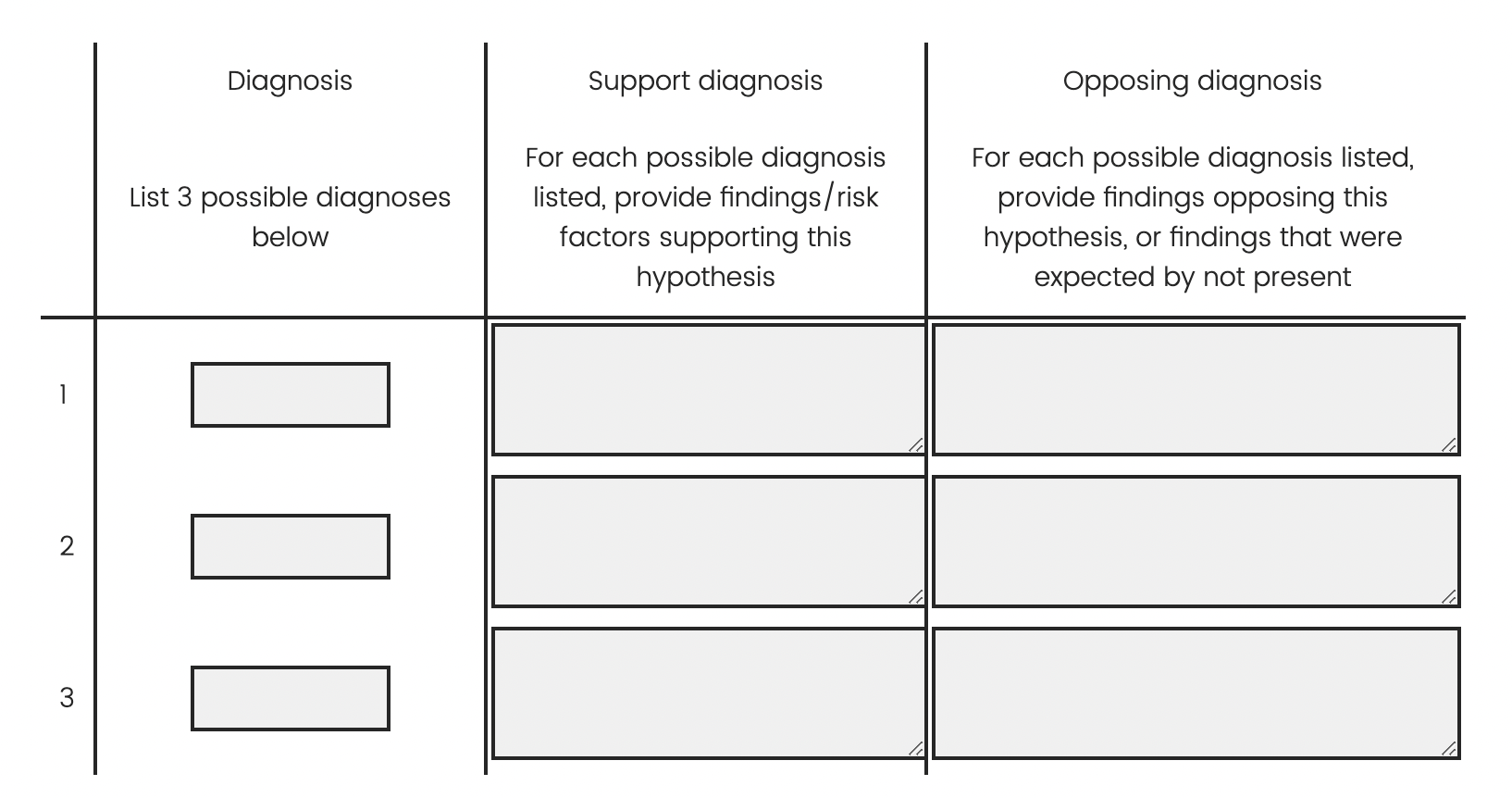 | |
| **PART 2 – Final diagnostic decision** | |
| **Question** | Based upon your reasoning above, what is your final diagnosis? |
| **PART 3 – Additional Steps** | |
| **Question** | Name up to 3 additional steps that you would take in your diagnostic process |

**SUPPLEMENTARY 2: GRADING RUBRIC**

**eTable 2: Structured-reflection rubric for Diagnostic Case #1, with example of high-scoring vs. low-scoring example responses**

**High Scoring Response**

| **Part 1 – Structured Reasoning** | | | |
| --- | --- | --- | --- |
| **Questions** | | **High Scoring Example** | **Scores** |
| **Question 1** | Diagnosis - List 3 Possible Diagnosis | Cholesterol embolism | 1/1 |
|  |  | Acute interstitial nephritis | 1/1 |
|  |  | Peripheral arterial disease | 1/1 |
| **Question 2** | Support Diagnosis - For each possible diagnosis listed, provide findings/risk factors supporting this hypothesis | Cholesterol embolism - Recent PCI, multi organ involvement (peripheral vasculature, renal, skin), rash, renal failure, eosinophilia, known CAD | 2/2 |
|  |  | Acute interstitial nephritis - Renal failure, rash, +/- anemia, proteinuria, all after intervention with many medications (including ASA) | 2/2 |
|  |  | Peripheral arterial disease - Leg pain associated with ambulation, improves with rest | 2/2 |
| **Question 3** | Opposing Diagnosis - For each possible diagnosis listed, provide findings opposing this hypothesis, or findings that were expected by not present | Cholesterol embolism - Would expect leg pain to be at rest (acute occlusion rather than more of a stable angina type picture), not sure would present with fever, anemia not necessarily explained either | 2/2 |
|  |  | Acute interstitial nephritis - Does not explain leg pain, did not receive classic causative medications / no other risk factors | 2/2 |
|  |  | Peripheral arterial disease - Would not expect nearly any of the lab abnormalities, unlikely to present this acutely | 2/2 |
| **PART 2 – Final diagnostic decision** | | | |
| **Question** | Based upon your reasoning above, what is your final diagnosis? | Cholesterol embolism | 2/2 |
| **PART 3 – Additional Steps** | | | |
| **Question** | Name up to 3 additional steps that you would take in your diagnostic process | Arterial ultrasounds of bilateral lower extremities / ABI, analyze urine for casts / eosinophils, nephrology consultation and consideration of renal biopsy | 2/2 |
| **Total score** | | | 18/19 |

*Caption:* *An example of a high-scoring response. The participant has three plausible diagnoses, with entirely correct and thorough answers for factors both supporting and opposing their diagnoses, as well as an entirely correct final diagnosis (cholesterol embolism).*

**Low Scoring Response**

| **Part 1 – Structured Reasoning** | | | |
| --- | --- | --- | --- |
| **Questions** | | **High Scoring Example** | **Scores** |
| **Question 1** | Diagnosis - List 3 Possible Diagnosis | Interstitial nephritis | 1/1 |
|  |  | Contrast-induced nephropathy | 0/1 |
|  |  | Pyelonephritis | 0/1 |
| **Question 2** | Support Diagnosis - For each possible diagnosis listed, provide findings/risk factors supporting this hypothesis | renal failure, eosinophilia, fever, rash | 2/2 |
|  |  | contrast exposure, acute renal failure | 2/2 |
|  |  | fever, aki | 1/2 |
| **Question 3** | Opposing Diagnosis - For each possible diagnosis listed, provide findings opposing this hypothesis, or findings that were expected by not present | Elevated GGT | 0/2 |
|  |  | rash, fever | 1/2 |
|  |  | rash, unremarkable ua | 1/2 |
| **PART 2 – Final diagnostic decision** | | | |
| **Question** | Based upon your reasoning above, what is your final diagnosis? | interstitial nephritis | 0/2 |
| **PART 3 – Additional Steps** | | | |
| **Question** | Name up to 3 additional steps that you would take in your diagnostic process | Renal ultrasound, urine eosinophils, urine sodium/creatinine ratio | 1/2 |
| **Total score** | | | 9/18 |

*Caption: An example of a low scoring response. The participant has only one plausible diagnosis for the case and has examples of incorrect or not fully correct for factors supporting and opposing their diagnoses. Their final diagnosis is incorrect, though their workup for their listed diagnoses is partially correct.*

**SUPPLENTARY 3:** **DISTRIBUTION OF DIAGNOSTIC PERFORMANCE SCORES**

**eTable 3: Distribution of Diagnostic Performance Scores of Physician + GPT-4 vs. Physician + Conventional Resources Only**

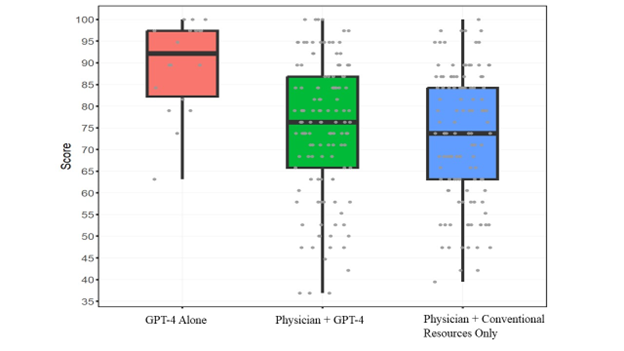

*Caption: Participants were randomized to access GPT-4 in addition to conventional diagnostic resources or to just conventional resources. GPT-4 alone scored 15.5 percentage points (95% CI 1.5 to 29, p=0.03) higher than the conventional resources group.*

**SUPPLENTARY 4: GPT-4 PERFORMANCE**

**eTable 4: GPT-4 Performance**

|  | **Overall** | **AI** | **Non-AI** | **Odds Ratio**  **(from mixed effect model)** |
| --- | --- | --- | --- | --- |
| **Final Diagnosis (Ordinal Scale)** |  |  |  | 1.4 [0.67 to 2.8],  p=0.39 |
| 0 | 80 (33%) | 36 (29%) | 44 (37%) |  |
| 1 | 7 (2.9%) | 6 (4.8%) | 1 (0.8%) |  |
| 2 | 157 (64%) | 83 (66%) | 74 (62%) |  |
| **Final Diagnosis (Binary)** |  |  |  | 1.9 [0.9 to 4.0],  p=0.10 |
| Incorrect | 80 (33%) | 36 (29%) | 44 (37%) |  |
| At least partially correct | 164 (67%) | 89 (71%) | 75 (63%) |  |

**SUPPLENTARY 5:** **GPT PROMPT AND RESPONSES**

**eTable 5: GPT Prompt and Responses for Diagnostic Case 1**

| **GPT-4 Prompt** | **GPT-4 Responses** |
| --- | --- |
| **Part 1 – Structured Reasoning**  You are an expert internal medicine physician solving a complex medical case for a test. You are going to receive a case vignette. After reading the case, I want you to give three parts of information.  Here is the first part:  First, you will list three possible diagnoses. Please number these 1., 2., 3.  Next, for each of the three diagnoses listed, you will provide the findings or risk factors that support this hypothesis.  Next, for each of the three diagnoses listed, you will provide findings opposing this hypothesis, or findings that were expected but not present.  **Part 2 – Final diagnostic decision**  Here is the second part: Please answer this question: Based upon your reasoning above, what is your final diagnosis?  **Part 3 – Additional Steps**  Here is the third part: Name up to 3 additional steps you would take in your diagnostic process.  **Diagnostic Case**  Here is the case:  History Of Present Illness  A 76M comes to his PCP complaining of pain in his back and thighs for 2 weeks. He has no pain sitting or lying, but walking causes severe pain in his low back, buttocks and calves. He feels febrile and tired. He was told by the referring cardiologist that his recent tests results since the pain started showed a new anemic and azotemia. A few days before the onset of the pain he had undergone coronary angioplasty. Heparin was administered for 48 hours.  Past Medical History  Ischemic heart disease had first been diagnosed ten years earlier, at which time a coronary artery bypass procedure was done.  Physical Examination  VITALS: 99.6° F.; pulse was 94/min and regular; BP was 110/88 mmHg.  GEN: Well appearing  CARDS: There is a grade III/VI apical systolic murmur; lungs were clear to auscultation.  PULM: Lungs are clear to auscultation bilaterally, no wheezing, or consolidations noted  ABD: Soft, non-tender to palpation  MSK: He does not have tenderness of his spine or pelvis. Spinal mobility is normal, as is the mobility of his hips. Standing is painless; however, pain is experienced in his low back, buttocks and calves within a minute of feeble running in place. The pain disappears shortly after exercise is discontinued.  EXT: Peripheral pulses were symmetrically reduced, but palpable. The neurological examination was normal.  SKIN: Patient has a purple, red, lacy rash over his low back and buttocks.  Laboratory  WBC of 11.5 x 103 cells /μL; differential of 64% segs, 20% lymphocytes, 3% monocytes, 12% eosinophils and 1% basophil. The hematocrit was 28% and the platelet count was 315 x 103 /μL. The erythrocyte sedimentation rate was 99 mm/h. Urinalysis was normal except for 2+ proteinuria. Serum creatinine was 4.0 mg/dL; sodium was 145 mEq/L; potassium 4.0 mEq/L; chloride 105 mEq/L. SGOT was 27 U/L; GGT was 90 U/L; alkaline phosphatase was 153 U/L. | Based on the provided medical case vignette, here are the three possible diagnoses along with supportive and opposing findings:  **Cholesterol Embolization Syndrome (CES)**  **● Supportive Findings:**   - Recent history of coronary angioplasty with heparin administration. - New onset anemia and azotemia post-procedure. - Purple, red, lacy rash over low back and buttocks (livedo reticularis). - Elevated erythrocyte sedimentation rate (ESR) and proteinuria.   ● **Opposing Findings:**   - Lack of findings typically associated with CES, such as renal failure, gastrointestinal symptoms, or neurological deficits.   **Peripheral Artery Disease (PAD)**  ● **Supportive Findings:**   - Pain in the lower back, buttocks, and calves that worsens with exercise (claudication). - History of ischemic heart disease, suggesting atherosclerotic disease elsewhere. - Symmetrically reduced peripheral pulses.   **● Opposing Findings:**   - Presence of a rash and high ESR are not typical features of PAD. - The rapid resolution of pain after stopping exercise is atypical for PAD.   **Vasculitis**  ● **Supportive Findings:**   - Presence of a rash that could be indicative of vasculitis. - Elevated ESR and eosinophilia. - Proteinuria and elevated creatinine suggesting renal involvement.   ● **Opposing Findings:**   - Lack of other systemic symptoms typically seen in vasculitis such as fever, weight loss, or joint pain.   **Final Diagnosis:**  Cholesterol Embolization Syndrome (CES) seems to be the most likely diagnosis. The recent angioplasty procedure, onset of anemia and azotemia, characteristic rash, and laboratory findings support this diagnosis more than the other possibilities.  **Further Diagnostic Steps:**   1. Skin Biopsy: To confirm the diagnosis of CES by identifying cholesterol crystals in small blood vessels. 2. Renal Biopsy: If renal involvement is suspected to confirm the presence of cholesterol emboli. 3. Vascular Imaging: Such as Doppler ultrasound or angiography to assess the extent of atherosclerotic disease and possible sources of cholesterol emboli. |

**SUPPLEMENTARY 6: ASSESSMENT TOOL VALIDATION**

**eTable 6: Assessment Tool Validation**

| **Item** | **Variance** |
| --- | --- |
| Diagnoses | 0.030 |
| Support diagnosis | 0.031 |
| Opposing diagnosis | 0.070 |
| Final diagnosis | 0.190 |
| Next steps | 0.042 |
